# Intensity of habitual physical activity and maintenance of normal blood pressure – findings from the SUN longitudinal cohort study

**DOI:** 10.1101/2024.04.29.24306595

**Authors:** Anne Katherine Gribble, Maria S. Hershey, José Francisco López-Gil, Fan-Yun Lan, Stefanos N. Kales, Miguel Ángel Martínez-González, Maira Bes-Rastrollo, Alejandro Fernandez-Montero

**Author notes:** **Corresponding author:** Alejandro Fernandez-Montero. Email address Department of Occupational Medicine, University of Navarra Clinic. Av. Pio XII, 36. 31008. Pamplona, Navarra, Spain.

## Abstract

(2)

**Background:** Physical activity (PA) is a modifiable protective factor against hypertension, but the optimum intensity of PA for prevention of hypertension remains unknown. It has been suggested that total energy expenditure is the crucial factor while intensity is non-differential provided it is moderate or above. Yet it is possible that higher intensity PA may produce a distinct effect.

**Methods:** We used data from the *Seguimiento Universidad de Navarra* (SUN) cohort – a large prospective longitudinal cohort study in Spain - to investigate how intensity of habitual PA may affect hypertension incidence. Average intensity of PA was calculated incorporating incidental walking and stairclimbing in addition to leisure-time PA (LTPA). Hazard ratios (HRs) for incident hypertension and their 95% confidence intervals (CI) were estimated using Cox regression analyses, and modelling adjusted for EE and body mass index (BMI) as well as other important covariables. Comparative models investigated how time spent in PA and EE in PA relate to hypertension incidence.

**Results:** 17,146 normotensive participants (63.6% female, mean age 36.7 years, mean BMI 23.2kg/m^2^) were followed for 204,677 person-years. 2,495 cases of incident hypertension emerged. After adjustment for covariables, including EE, intensity of PA was monotonically associated with decreased risk for incident hypertension (aHR for Q5 vs Q1: 0.81, 95% CI 0.71-0.93). In comparison, increasing time in PA was associated with increasing risk for incident hypertension following adjustment for EE (aHR for Q5 vs Q1: 1.60, 95% CI 1.10- 2.32).

**Conclusion:** Intensity of habitual PA is independently and inversely associated with incidence of hypertension.

## INTRODUCTION

Hypertension is a leading modifiable risk factor for cardiovascular disease and death.^1–3^ Although medical management of hypertension may mitigate morbidity and mortality, it is associated with significant accumulative health care burden and cost.^4,5^ Primary prevention must be a priority.

Physical activity (PA) is a modifiable protective factor known to aid in the maintenance of normal blood pressure (BP).^6–11^ Given the scope for modification of PA, investigation into what modifications of PA may lead to most protective effect is particularly valuable.

Most prior studies quantify and analyse PA in terms of energy expenditure (EE).^10,11^ However, modifiable factors such as intensity, duration, frequency and type of PA also merit attention.^8,9,12^ The intensity of PA, measured as the metabolic equivalent of a task (MET), is a key determinant of EE. Multiple previous studies found that more intense PA was more protective against cardiovascular disease and mortality than PA at lower intensity, however, they were unable to conclude whether the benefit from intensity was due to an associated increase in EE or due to an effect of higher intensity itself.^13^ Hence, the current recommendations from the World Health Organisation and other leading groups reflect the dominant view that moderate intensity and vigorous intensity exercise have equal benefit provided that EE is also equal.^14–16^ But it is also possible that the intensity of PA may produce an effect independent of its effect on EE. Evidence from the *Seguimiento Universidad de Navarra* (SUN) cohort has shown that, even after controlling for EE, higher intensity of PA is associated with reduction in incidence of cardiovascular disease,^17^ metabolic syndrome,^18^ and type 2 diabetes^19^ and also that faster self-perceived walking pace is associated with lower rates of hypertension.^20^ The aim of this study was to examine in the same cohort the effect of habitual intensity of PA on hypertension incidence.

## METHODS

### Study population

The *Seguimiento Universidad de Navarra* (SUN) study is a multipurpose longitudinal cohort study focused on Mediterranean diet and lifestyle patterns in relation to cardiovascular and other outcomes.^21^ It follows a long-running dynamic cohort, permanently open since December 1999. Participants are graduates of Spanish universities and over 50% hold health-related professional degrees. At entry, a baseline questionnaire collects detailed information on diet, lifestyle and health-related characteristics. Follow-up questionnaires are distributed every 2 years and can be completed online or on hardcopy. Data is self-reported. Voluntary completion and return of the baseline questionnaire is taken as informed consent in accordance with the Declaration of Helsinki.^22^ The study’s protocol was approved by the Institutional Review Board of the University of Navarra and registered at clinicaltrials.gov (NCT02669602).

The SUN database included 23,133 participants as of May 2022. Those without follow-up were excluded from the analysis, as too were those whose self-reported data was deemed unreliable because their responses to the food frequency questionnaire resulted in energy intake totals outside a plausible range.^23^ Participants with pre-existing diagnosed hypertension were excluded, as were those already on anti-hypertensives (beta-blockers, calcium channel blockers, angiotensin-converting-enzyme inhibitors and angiotensin receptor II blockers) and those with pre-existing hypertensive range blood pressures (systolic BP ≥140mmHg or diastolic BP ≥ 90mmHg).^24^ There remained 17,146 participants available for inclusion (Figure 1).

**Figure 1.**
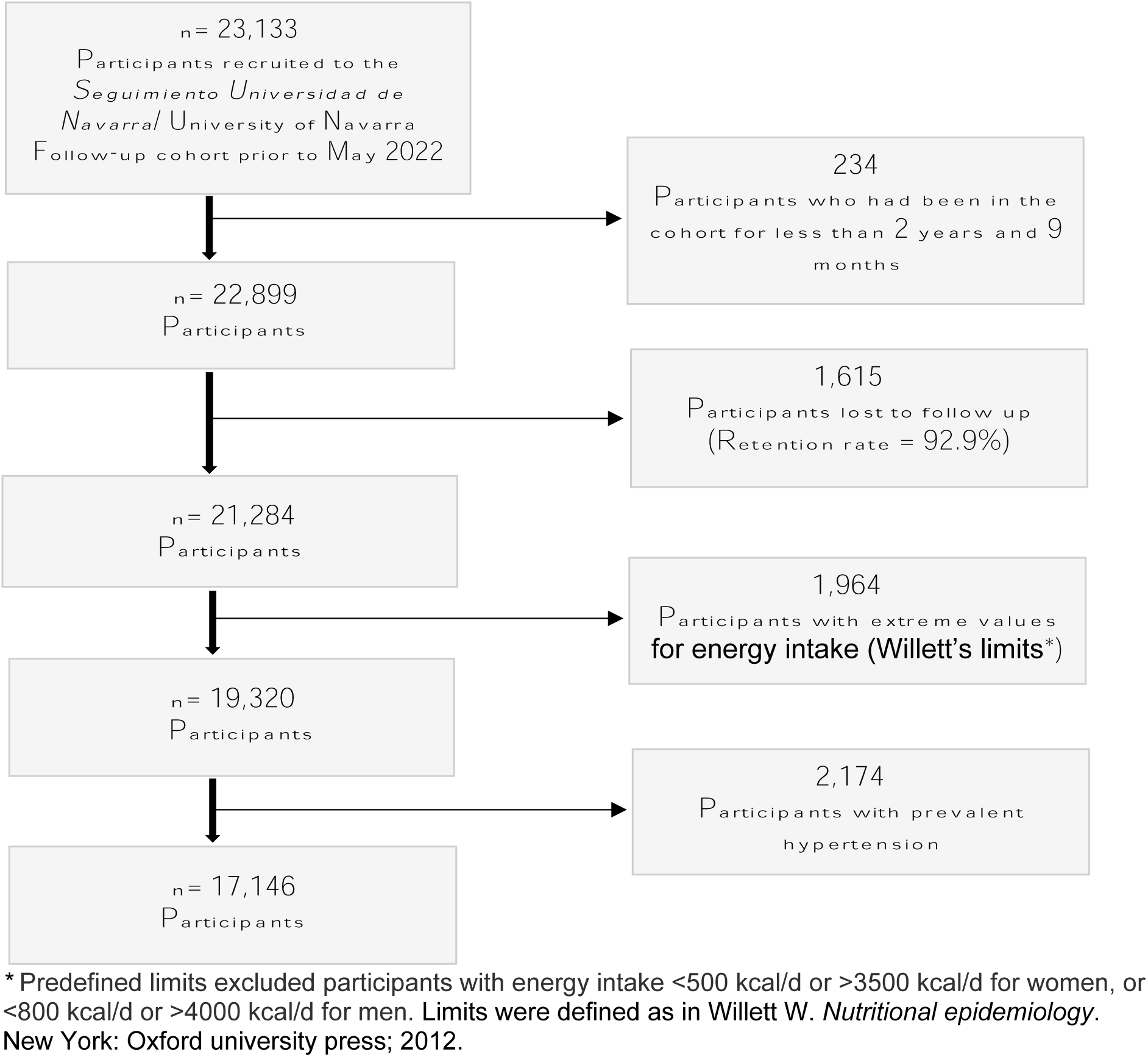
Flow chart of study sample selection from the Seguimiento Universidad de Navarra/University of Navarra Follow-up cohort. 1999-2022.

### Exposure assessment – Intensity of physical activity

Intensity of PA for each participant was calculated based on self-estimated usual engagement in PA and included incidental exercise (PA resulting from activities of daily living) and leisure-time physical activity (LTPA).

The questions to assess incidental PA asked, “How much do you usually walk each day?” (answer choices: <10 mins/10-20 mins/21-30 mins/30 mins-1hr/1-2 hrs/>2 hrs) and “How many flights of stairs do you climb each day?” (≤2/3-4/5-9/10-14/ ≥15). Flights of stairs were converted into an estimated time (≤2 flights = 30s, 3-4 flights = 1min 45s, 5-9 flights = 3 mins 30s, 10-14 flights = 6 mins, ≥15 flights = 8 mins 45s). 225 participants (1.3% of the study sample) were missing data for stair-climbing and only 139 participants (0.8% of the study sample) were missing data for incidental walking. These missing values were predicted using logistic regression with age and sex as explanatory variables.

Time spent in LTPA was assessed taking seasonality into account. Participants answered “Do you exercise?” (Yes/No), then proceeded to a 17-item LTPA question set entitled “During the last year, how much time on average did you spend doing the following activities?”. Listed activities were “walking”; “jogging”; “cycling”; “stationary bike”; “racket sports”; “soccer and indoor soccer”; “other team sports (e.g., basketball, handball)”; “dance and aerobics”; “hiking and rock-climbing”; “gym-based work outs”: “gardening, yard maintenance, pool maintenance and renovations”; “skiing, ice-skating and rollerblading”; “judo, karate and martial arts”; “sailing” and “other sports or activities not listed”. For each activity, participants answered “Average time per week” (Never/1-4 mins/5-19 mins/20-59 mins/<1hr/1-1.5hrs/2-3hrs/4-6 hrs/7-10hrs/≥11hrs) and “Months per year” (<3 months/3-6 months/>6 months.) This question set has been validated in our cohort.^25^ Reported time per week was multiplied by 0.125 for <3 months, or 0.375 for 3-6 months, or 0.75 for >6 months. Complete non-response for any activity was interpreted as non-participation. In contrast, if either “hours per week” or “months per year” was answered but not both, the missing value was predicted by logistic regression using age and sex as explanatory variables. As many as 6445 participants (37.6% of the sample) had at least one missing value to be predicted, whilst 854 participants (5.0%) required prediction of more than five values.

METs for each activity were drawn from the Compendium of Physical Activities.^26,27^ METs for walking depended on self-report of walking pace (slow/average/fast/very fast). 159 participants (0.9% of the sample) missing values for walking pace were assigned “average” pace.

Total weekly exercise time and total weekly energy expenditure (EE) were calculated. Then, average intensity of PA was calculated by dividing EE by exercise time. Quintiles of intensity were used as categories for analysis.

### Outcome assessment – Incident hypertension

Incidence of hypertension was assessed based on participant report of hypertension diagnosis by a physician. Follow-up questionnaires asked if participants had been diagnosed with hypertension, defined in-line as systolic BP>130mmHg or diastolic BP>85mmHg, and their month and year of diagnosis. This assessment of this outcome has been validated in a subsample of our cohort.^28^ Participants’ regular medications were also recorded at each point of follow-up, so a sensitivity analysis was performed counting participants taking antihypertensive medications as additional positive cases.

### Covariables

Covariables for adjustment were sex (male / female), age (years), calendar year of completion of first questionnaire, years of tertiary education (years), body mass index (BMI) (kg/m^2^), total energy intake (kcal/day), Mediterranean Diet Score (Trichopoulous score)^29^, special diets (yes / no), sodium intake (mg/day), alcohol intake (g/day), coffee intake (cups/day), smoking exposure (pack years), television viewing time (hours/day), night-time sleep (hours/night), cardiovascular disease (yes/no), diabetes (yes/no), cancer (yes/no), parental history of hypertension (yes/no). Television viewing time and night-time sleep were the only covariables with missing values. Regression-base simple imputation involving all non-missing covariables was used to impute these missing values. Continuous covariables were reduced to quintiles for the analysis, except age (divided in decades) and calendar year of first questionnaire (divided in 6 groups).

### Statistical analysis

Cox regression analyses were used to study the relationship between intensity of PA and time to hypertension incidence. The end point was whichever occurred first out of hypertension diagnosis, death or last completed follow-up questionnaire. The Breslow method was used for tied observations. The lowest quintile of intensity was the reference for comparison. We performed an unadjusted analysis, then an analysis adjusting for sex and stratifying for age and year of cohort entry, and, finally, a multivariable analysis adjusting for EE as well as all other covariables with stratification for age and year of cohort entry.

To enable comparison between PA intensity and other PA variables, the relationship between time in PA and hypertension incidence and that between EE and hypertension incidence were also examined using the same series of Cox regression models as that described above, with the exception that the multivariable-adjusted Cox regression model for EE did not include EE as additional adjustment variable.

To consider potential non-linearity in the relationships between PA factors and incident hypertension, cubic spline regression models were fitted exploring PA intensity and time in PA as continuous variables.

To compare the effects of intensity versus time in PA, nine categories were created based on intersection tertiles of PA intensity and time in PA and analysed using multivariable Cox regression (no adjusting for EE) with the intersection of the lowest terciles of PA intensity and PA time as referent category.

Stratified Cox regression analysis was used to examine subgroups of age (<50 years vs ≥50 years), sex (male vs female), overweight (<25kg/m^2^ vs ≥25kg/m^2^), parental history of hypertension (yes vs no), hours of television viewing (above or below sample median) and amount of night-time sleep (recommended 7-9 hours^30^ vs other). We applied the likelihood ratio test of nested models to check for interaction by these a priori selected factors.

We performed multiple sensitivity analyses: one excluding participants with missing data for PA variables; another excluding participants with incidence of hypertension before two years of follow up or with follow-up time less than two years; another declaring maximum end point at 12 years follow-up time; another including additional cases of incident hypertension, identified based on newly reported use of hypertensive medications or self-report of systolic BP >140mmHg or diastolic BP >90mmHg (diagnosis based on self-reported in-office BP has been previously validated in our cohort).

## RESULTS

Of the 17,146 participants included in the analysis, 10,910 (63.6%) were women and 9,569 (55.8%) were health professionals. The mean age at cohort entry was 36.7 years and the mean BMI was 23.2kg/m^2^. During follow-up (204,677 person years), there emerged 2,495 cases of incident hypertension, equating to an absolute risk of 14.55%.

Median intensity of habitual PA was 4.1 METs (IQR 3.8 to 4.8). Median weekly time in PA was 5.0 hours (IQR 2.5 to 8.8). Median EE was 21.4 MET-hrs per week (IQR 10.6 to 38.4). On average, incidental exercise accounted for 54.3% of EE and 57.4% of time in PA across the week. Walking was also the most common LTPA, with 70.9% of participants engaging in walking for leisure.

The hazard of hypertension increased with age: the probability of being hypertension free at 40 years old was 91% (95% CI 90 to 92), whereas the probability of being hypertension free at 80 years old reduced to 33% (95% CI 30-35). Mean age at time of hypertension diagnosis was 51.7 years (95% CI 51.2 to 52.2) and median follow-up time prior to diagnosis was 5.9 years (IQR 3.4 to 11.4). Ten percent of cases reported diagnosis within 1.6 years of cohort entry. As this raised the possibility of inverse causation, a sensitivity analysis was performed excluding all participants with hypertension incidence before 2 years follow-up.

The baseline characteristics of our study sample across quintiles of PA intensity are presented in Table 1. Women were under-represented in the uppermost quintile of PA intensity, but participant profiles otherwise differed minimally across intensity quintiles.

**Table 1.** Baseline characteristics of study sample according to quintiles of average intensity of physical activity.

|  | Quintiles of average intensity of physical activity (metabolic equivalents) |  |  |  |  |
| --- | --- | --- | --- | --- | --- |
|  | Q1<br>(2.5-3.6 METs) | Q2<br>(3.6-4.0 METs) | Q3<br>(4.0-4.4 METs) | Q4<br>(4.4-4.9 METs) | Q5<br>(4.9-10.5 METs) |
| N | 3,430 | 3,496 | 3,362 | 3,429 | 3,429 |
| Women (%) | 69.45 | 71.14 | 68.53 | 63.14 | 45.84 |
| Age (yrs) | 39.4 (12.5) | 37.3 (11.9) | 36.1 (11.1) | 35.3 (10.4) | 35.3 (10.0) |
| Year of cohort entry | 2004 (3) | 2004 (4) | 2004 (4) | 2004 (4) | 2004 (4) |
| Years of university (years) | 4.9 (1.4) | 5.0 (1.4) | 5.0 (1.5) | 5.1 (1.5) | 5.2 (1.6) |
| Energy intake (kcal/day) | 2286 (618) | 2333 (594) | 2343 (601) | 2363 (612) | 2402 (622) |
| Special diets (%) | 7.58 | 7.18 | 7.53 | 6.79 | 7.61 |
| Mediterranean Diet Score (score out of 9) | 4.0 (1.7) | 4.2 (1.8) | 4.2 (1.8) | 4.2 (1.8) | 4.2 (1.8) |
| Sodium intake (mg/day) | 3261 (2063) | 3248 (2044) | 3313 (2152) | 3293 (1968) | 3499 (2716) |
| Coffee intake (cups/day) | 1.2 (1.3) | 1.2 (1.3) | 1.2 (1.2) | 1.3 (1.3) | 1.2 (1.3) |
| Alcohol intake (g/day) | 5.4 (9.0) | 5.9 (9.3) | 6.0 (8.5) | 6.5 (9.5) | 7.2 (9.1) |
| Smoking exposure (pack years) | 7.2 (10.8) | 6.0 (9.6) | 5.0 (8.8) | 4.7 (7.9) | 4.6 (7.9) |
| Night-time sleep (hours/night) | 7.4 (0.9) | 7.3 (0.9) | 7.4 (0.9) | 7.3 (0.9) | 7.3 (0.8) |
| Television (hours/day) | 1.8 (1.3) | 1.7 (1.3) | 1.6 (1.2) | 1.5 (1.2) | 1.5 (1.1) |
| Body mass index (kg/m<sup>2</sup>) | 24.0 (3.8) | 23.1 (3.4) | 22.8 (3.2) | 22.9 (3.1) | 23.2 (2.9) |
| Prevalent cardiovascular disease (%) | 0.96 | 0.83 | 0.74 | 0.55 | 0.70 |
| Prevalent diabetes (%) | 1.40 | 1.34 | 1.37 | 0.90 | 0.73 |
| Prevalent cancer (%) | 2.83 | 2.77 | 2.47 | 1.92 | 1.81 |
| Prevalent depression (%) | 12.94 | 11.21 | 10.95 | 10.12 | 9.30 |
| Parental history of hypertension (%) | 24.52 | 24.40 | 24.54 | 26.04 | 23.94 |
| Time spent in physical activity (hours/week) | 5.1 (4.0) | 6.0 (4.6) | 6.5 (4.8) | 6.6 (5.3) | 7.9 (6.2) |
| Energy expenditure in physical activity (MET-hrs/week) | 17.0 (13.5) | 23.1 (17.9) | 27.0 (20.3) | 30.4 (24.7) | 44.9 (36.4) |

*Continuous variables are expressed as mean with standard deviation in parentheses and categorical variables are expressed as percentages.*

Cox regression modelling (Table 2) revealed that higher intensity of PA was associated with reduced risk of incident hypertension in a monotonic pattern (*p* for trend = <0.001). The risk of incident hypertension was 19% less in the quintile of highest intensity when compared to the lowest. Similarly, increasing EE appeared to reduce risk of incident hypertension: after multivariable adjustment, the fourth and fifth quintiles for EE had relative risk reduction of 16% and 21% respectively compared to Q1. In contrast, increasing only weekly time in PA without significant change to EE was associated with increased hypertension incidence (adjusted-HR for Q5 vs Q1 = 1.60, 95% CI 1.10-2.32).

**Table 2.** Hazard ratios for the incidence of hypertension according to quintiles of physical activity parameters - intensity of physical activity, time spent in physical activity, and total energy expenditure in physical activity

| **Average intensity of physical activity (metabolic equivalents)** |  |  |  |  |  |  |
| --- | --- | --- | --- | --- | --- | --- |
|  | **Q1**<br>(2.5-3.6<br>METs) | **Q2**<br>(3.6-4.0<br>METs) | **Q3**<br>(4.0-4.4<br>METs) | **Q4**<br>(4.4-4.9<br>METs) | **Q5**<br>(4.9-10.5<br>METs) | P for<br>trend |
| Person-years | 39813.9 | 41871.4 | 40272.2 | 41350.5 | 41369.1 |  |
| Cases | 618 | 526 | 461 | 457 | 433 |  |
| Crude HR | 1 (Ref.) | 0.89 (0.79-1.00) | **0.86 (0.76-**<br>**0.97)** | 0.88 (0.78-1.00) | **0.82 (0.72-**<br>**0.92)** | 0.002 |
| Age and sex<br>adjusted HR | 1 (Ref.) | 0.89 (0.79-1.00) | **0.84 (0.74-**<br>**0.95)** | **0.83 (0.74-**<br>**0.94)** | **0.71 (0.62-**<br>**0.80)** | <0.001 |
| Multivariable<br>adjusted HR*<sup>§</sup> | 1 (Ref.) | 0.95 (0.84-1.07) | 0.94 (0.83-1.06) | 0.93 (0.82-1.06) | **0.81 (0.71-**<br>**0.93)** | 0.004 |
| **Time spent in physical activity per week (hours)** |  |  |  |  |  |  |
|  | **Q1**<br>(0.8-2.0hrs) | **Q2**<br>(2.0-3.9hrs) | **Q3**<br>(3.9-6.4hrs) | **Q4**<br>(6.4-10.2<br>hrs) | **Q5**<br>(10.2-53.3<br>hrs) | P for<br>trend |
| Person-years | 40767.9 | 42051.1 | 39870.2 | 40900.9 | 41086.9 |  |
| Cases | 505 | 485 | 500 | 506 | 499 |  |
| Crude HR | 1 (Ref.) | **0.87 (0.76-**<br>**0.98)** | 0.92 (0.81-1.04) | 0.89 (0.79-1.01) | **0.85 (0.75-**<br>**0.96)** | 0.048 |
| Age and sex<br>adjusted HR | 1 (Ref.) | **0.84 (0.74-**<br>**0.95)** | **0.88 (0.78-**<br>**0.99)** | **0.84 (0.74-**<br>**0.95)** | **0.77 (0.68-**<br>**0.88)** | 0.001 |
| Multivariable<br>adjusted HR*<sup>§</sup> | 1 (Ref.) | 0.95 (0.75-1.21) | 1.28 (0.95-1.71) | **1.44 (1.03-**<br>**2.00)** | **1.60 (1.10-**<br>**2.32)** | 0.009 |
| **Total energy expenditure in physical activity per week (metabolic equivalent hours)** |  |  |  |  |  |  |
|  | **Q1**<br>(2.2-8.5<br>MET-hrs) | **Q2**<br>(8.5-16.3<br>MET-hrs) | **Q3**<br>(16.3-26.8<br>MET-hrs) | **Q4**<br>(26.8-43.7<br>MET-hrs) | **Q5**<br>(43.7-354.3<br>MET-hrs) | P for<br>trend |
| Person-years | 40143.1 | 40664.5 | 41380.0 | 41357.2 | 41132.3 |  |
| Cases | 508 | 509 | 514 | 486 | 478 |  |
| Crude HR | 1 (Ref.) | 0.91 (0.81-1.03) | 0.89 (0.79-1.01) | **0.84 (0.74-**<br>**0.95)** | **0.83 (0.73-**<br>**0.94)** | 0.004 |
| Age and sex<br>adjusted HR | 1 (Ref.) | 0.89 (0.79-1.01) | **0.85 (0.75-**<br>**0.96)** | **0.80 (0.70-**<br>**0.90)** | **0.73 (0.65-**<br>**0.83)** | <0.001 |
| Multivariable<br>adjusted HR* | 1 (Ref.) | 0.94 (0.83-1.07) | 0.89 (0.79-1.01) | **0.84 (0.74-**<br>**0.96)** | **0.79 (0.70-**<br>**0.90)** | <0.001 |

\* Adjusted for sex (male/female), age (years), calendar year of entry into the cohort, years of university (years), total energy intake (kcal/day), Mediterranean Diet Score (score out of 9), special diet (yes/no), sodium intake (mg/day), coffee intake (cups/day), alcohol intake (g/day), smoking exposure (pack years), night-time sleep (hours/night), television viewing time (hours/day), body mass index (kg/m<sup>2</sup>), prevalent cardiovascular disease (yes/no), prevalent diabetes mellitus (yes/no), prevalent cancer (yes/no), parental history of hypertension (yes/no), change in regular physical activity as reported during early follow up (no change/increased/decreased).
<sup>§</sup> Also adjusted for total energy expenditure (MET-hrs/week).

Regression models based on categorisation by intersecting tertiles of PA intensity and time in PA found that the risk reduction for the highest tertile of intensity was significant across the entire range of time in PA, whereas the risk reduction from maximising time in PA was not significant in combination with the lowest tertile of PA intensity (Figure 3).

**Figure 2.**
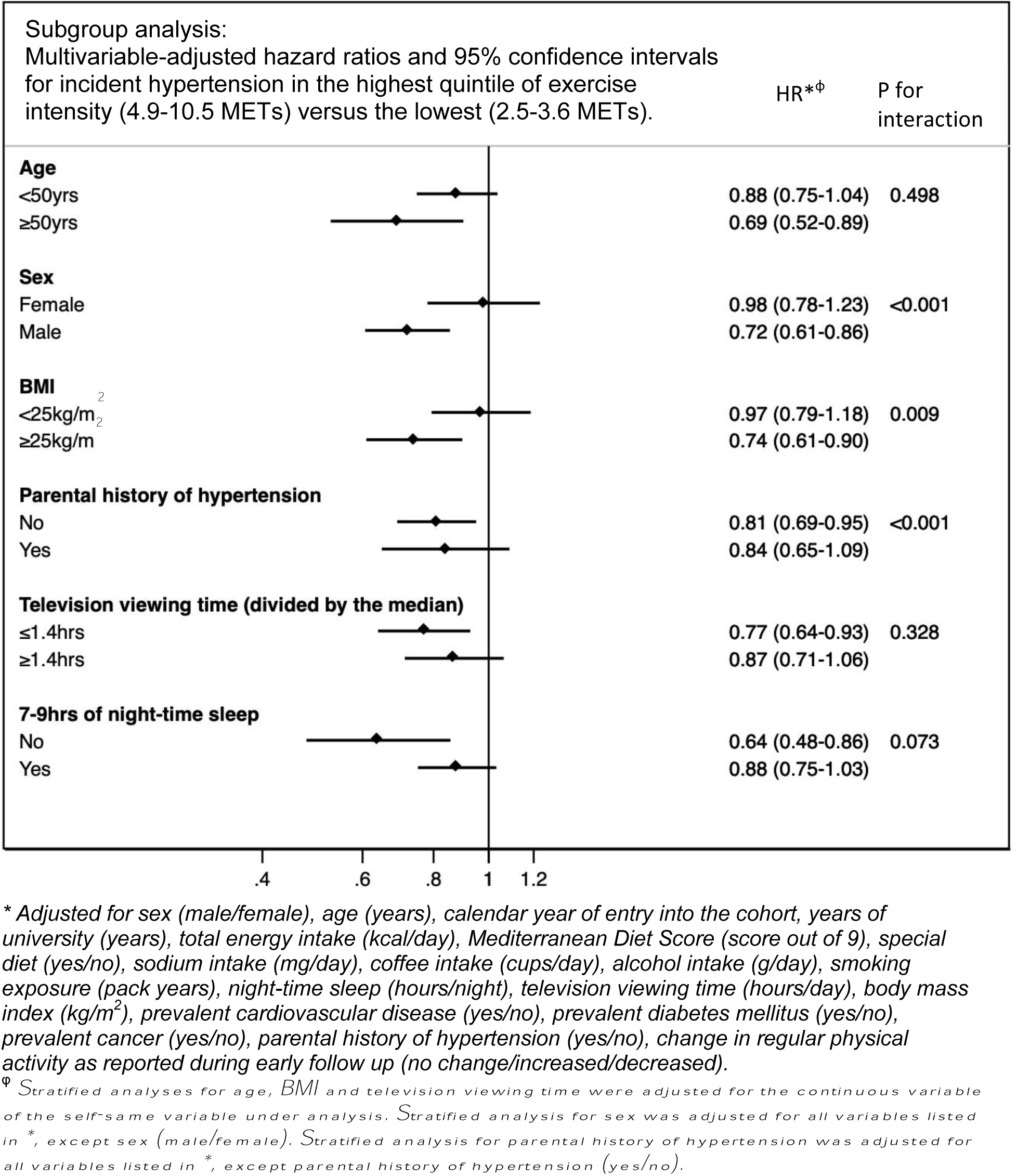
Subgroup analysis: Multivariable-adjusted hazard ratios and 95% confidence intervals for the incidence of hypertension in the highest quintile of exercise intensity (4.9-10.5 METs) versus the lowest (2.5-3.6 METs).

**Figure 3.**
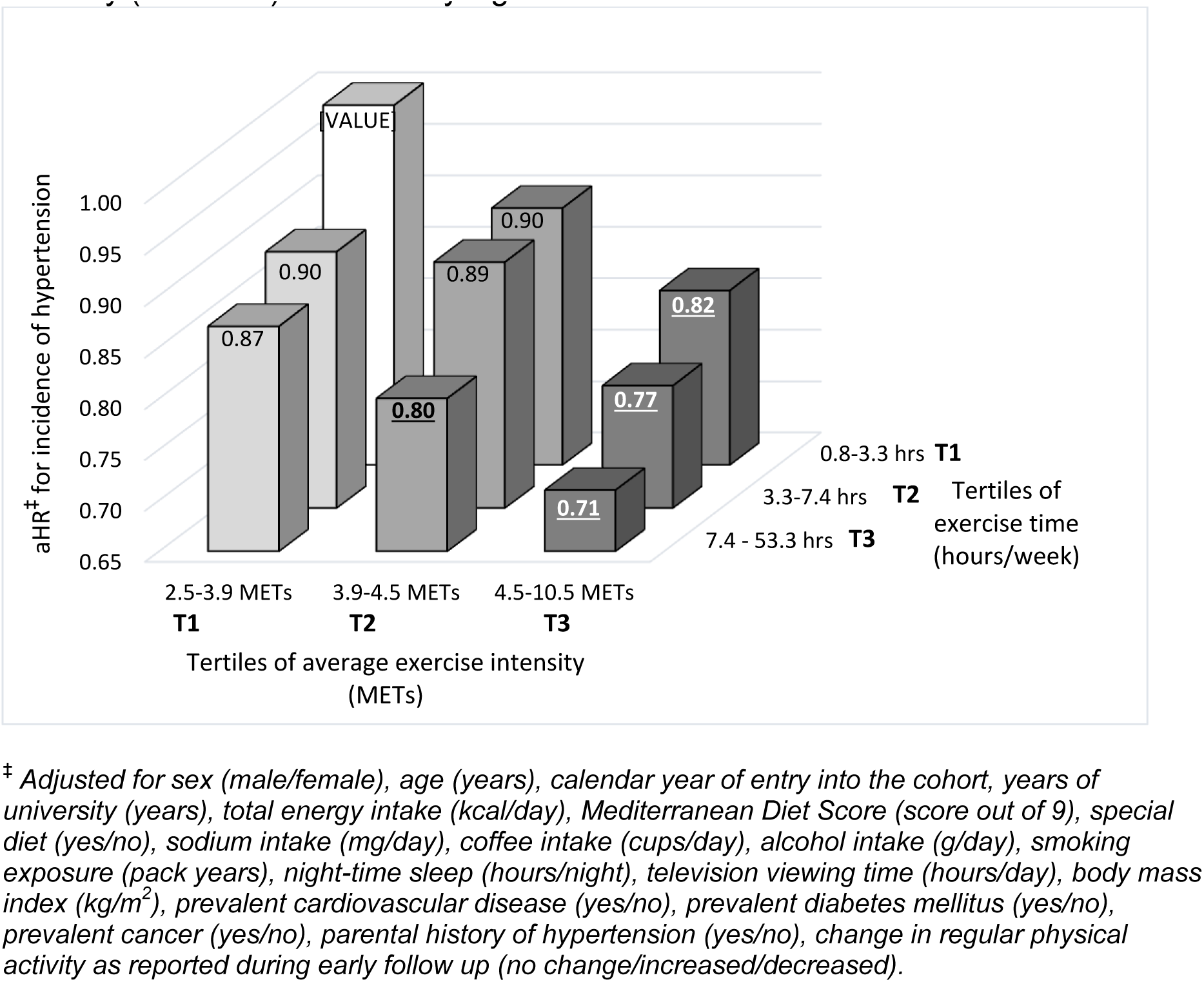
Multivariable adjusted^‡^ hazard ratios for the incidence of hypertension comparing the effect of weekly exercise time (in tertiles) and average exercise intensity (in tertiles). Statistically significant results marked in bold and underlined.

Cubic spline regressions (Figure 4) revealed that the inverse association between hazard of incident hypertension and intensity of PA and strengthened at a relatively constant gradient across the continuum of intensity. In contrast, the relationship between incident hypertension and weekly time in PA was not linear: from 2-5 hours per week, each additional hour strengthened the inverse association, but subsequent increase from 5 to 10 hours brought almost no additional effect and above 10 hours of weekly PA, the change per each additional hour was small.

**Figure 4.**
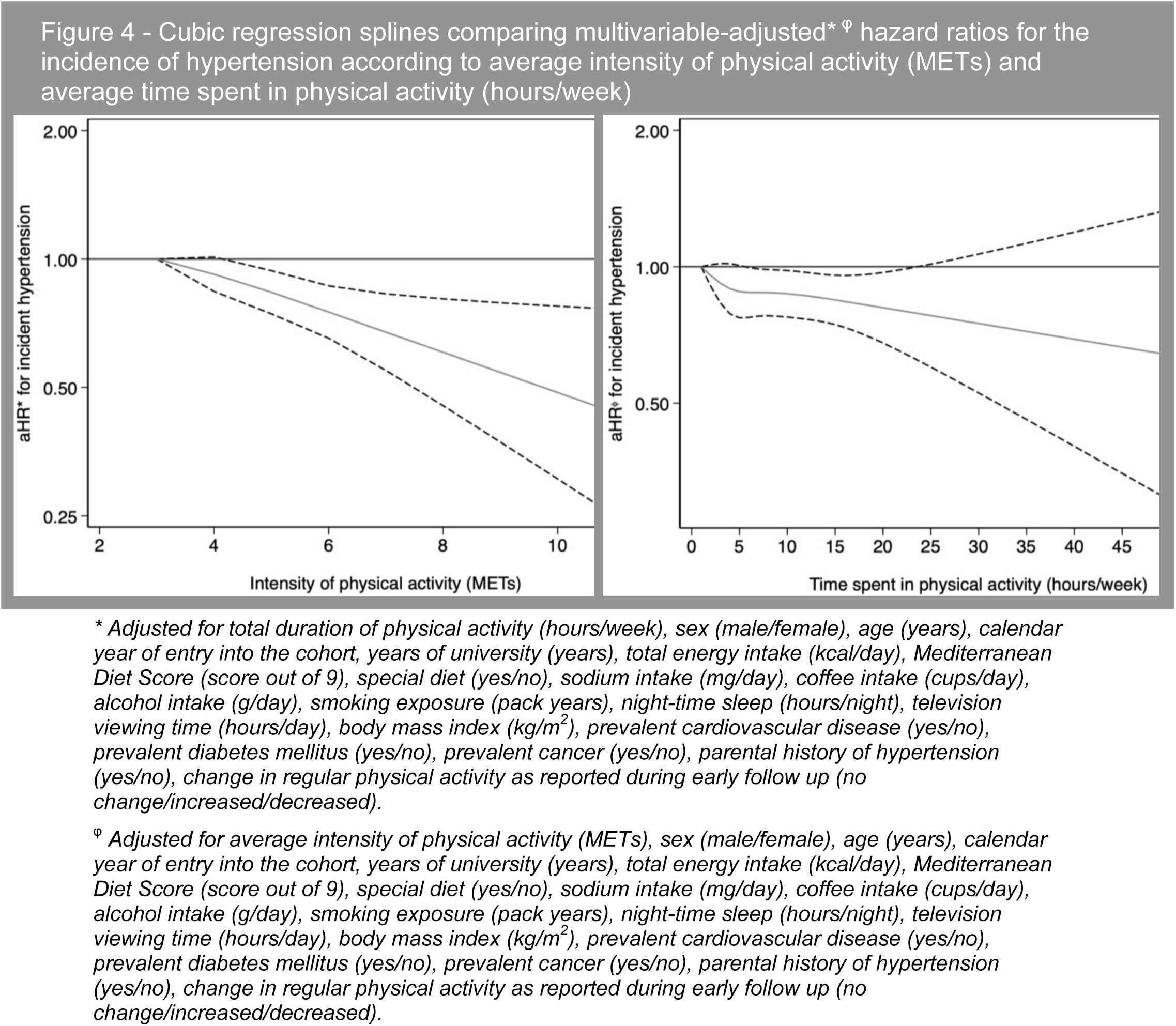
– Cubic regression splines comparing multivariable-adjusted* ^φ^ hazard ratios for the incidence of hypertension according to average intensity of physical activity and average weekly time performing physical activity (hours/week)

Subgroup analysis found that the risk reduction attributed to the upper quintile of PA intensity was greater for participants who were older, male, overweight, those who spent less time watching TV and those who did not sleep the recommended 7-9 hours each night. Sex and overweight were identified as interaction variables affecting the relationship between PA intensity and risk of hypertension incidence.

## DISCUSSION

This study found that as habitual PA intensity increased, incident hypertension decreased, even after controlling for EE. To identify the independent effect of intensity any analysis must control for the effect of EE,^31^ whilst also noting the trade-off whereby if intensity increases without increasing EE, then time spent in PA necessarily decreases. Although leading guidelines advise that maximum health benefit from PA can be obtained by increasing time in PA to increase EE,^14–16^ our findings suggest that to reduce risk of incident hypertension one should instead prioritise increasing PA intensity.

This finding constitutes a relatively new perspective as compared to previous studies into the relationship between PA and incident HTN. Whilst it is well established that greater EE in PA reduces the risk of developing hypertension,^8,10,12,32^ there is little clarity regarding targets for intensity. Many interventional studies provide evidence that moderate intensity exercise protects against incident hypertension,^10^ however most of these studies only compare moderate intensity PA against a reference group engaging in little or no PA, not against a higher intensity.^10^ Thus, their findings could point to a dose-response relationship between intensity of PA and prevention of incident hypertension, rather than the superiority of moderate level intensity. There are few interventional studies in normotensive subjects examining different intensities of PA in relation to incident hypertension or change in ambulatory BP,^12,31^ possibly because randomisation to high intensity PA interventions may lead to safety concerns or drop-out bias. The evidence from these few interventional studies possibly suggests that the greater the intensity of PA the greater the reduction in BP, ^31^ but the strength of the evidence is severely limited by the small samples and short follow-up. A Cochrane review entitled “Walking for Hypertension” estimated a 3.68mmHg reduction in systolic BP following walking interventions in normotensives,^33^ whilst a systematic review and meta-analysis of running interventions estimated that systolic BP was reduced by 4.2mmHg following running interventions in normotensive populations.^34^ Thus, interventional studies currently provide no clear evidence of clinically significant difference in effect of moderate versus higher intensity PA on BP.

The findings from three longitudinal cohort studies are similarly suggestive but inconclusive. Paffenbarger’s study of 14,998 male Harvard alumni found that vigorous sports reduced risk of incident hypertension, such that alumni who did not play vigorous sports were 35% more likely to develop hypertension after 6-10 years of follow up.^35^ However, the quality of the evidence is limited because the study did not characterise the amount of vigorous sports and had minimal adjustment for relevant confounders.

Another study compared the National Walkers Cohort against the National Runners Cohort to elicit a comparison between moderate versus high intensity PA.^36^ This study found runners were less likely than walkers to develop hypertension, however since the analysis did not control for EE, the authors reasoned that the difference owed to runners’ additional EE in PA rather than increased intensity per se. Nonetheless, when they then compared walkers and runners with equal EE in an analysis adjusting for BMI, the risk reduction from running surpassed that from walking. They also found that faster pace in both cohorts was associated with reduced risk of in hypertension, though again they concluded that this difference was due to faster pace increasing EE and not due to an independent effect of increased intensity. In any case, the comparison of intensity was limited because average intensity was not calculated and participants in both cohorts reported engaging in mainly vigorous intensity PA outside of their nominal walking or running.

A final study followed 11,285 participants from the Australian Women’s Longitudinal Study and examined the effect of moderate versus moderate-high intensity LTPA on BP.^37^ This study was able to adjust for EE and found that at all levels of EE, women who engaged in moderate-vigorous PA had lower risk of hypertension than those who engaged in moderate intensity PA alone. However, due to overlapping confidence intervals, they found no evidence of a difference between the two groups.

As such, our study is important because it provides evidence that increasing PA intensity reduces risk of incident hypertension, an association which has been hinted by previous studies at but never conclusively established.

Some of the biological pathways by which PA contributes to lower ambulatory BP are probably accentuated by increased intensity of PA.^38^ During exercise, heart rate, stroke volume and BP increase to meet the demands of working muscle. The higher the intensity, the greater the change in these cardiac parameters. The ability to take up, transport and use oxygen during exercise is known as cardiorespiratory fitness (CRF). CRF is conditioned by PA and best conditioned by training above a minimum intensity of 50% of VO2max.^13,39^ Increased CRF is associated with lower rates of hypertension.^40^ Regular high intensity PA causes exercise-induced cardiac remodelling (EICR).^41^ These structural adaptions, known as the ‘athlete’s heart’, include increased myocardial mass, enlargement of the four chambers, increased cardiac muscle contractility and improved diastolic function, changes which facilitate increased cardiac output during exercise as well as a lower heart rate at rest.^41^ Training also increase arterial compliance due to endothelial reaction to the change in stroke pressures.^38^ In keeping with this theory, athletes have been shown to have lower incidence of hypertension compared to age-matched healthy controls and athletes’ arteries have been found to be 30 years less stiff than the arteries of age-matched healthy controls.^42^

Our study is valuable because of its large sample size, low drop-out rate and the longitudinal study design. We were able to compare intensity within the same cohort and adjust for critical confounders such as EE and BMI. Moreover, our analysis of PA intensity, time in PA and EE reflects the totality of PA, including both LTPA and incidental PA. That said, our study is limited by its imprecise assessment of exercise intensity. The LTPA question set has been validated in our cohort, however participant responses may be affected by recall bias and the approximation of seasonality is crude. Our assessment of incidental PA components have yielded meaningful results in other studies within our cohort,^20^ but have not been validated. The Compendium of Physical Activities intensity values are the accepted standard for use in epidemiological studies, but they are generic and cannot reflect factors such as the participant’s CRF, their activity-specific fitness, or their approach – whether effortful or relaxed, factors which may significantly alter the true intensity of an activity. Future studies will be able to assess intensity more accurately, for example by using personal activity intelligence and wearable heart rate tracking devices.^43^

### Perspectives

In conclusion, this study of 17,146 participants provides evidence that comparatively higher intensity PA may be more protective against incident hypertension than comparatively lower intensity PA. Individuals should increase the average intensity of their habitual PA, either through adopting new higher intensity activities or by adapting low and moderate intensity activities to make them more effortful – for example, increasing habitual walking pace would increase average intensity even though walking may never meet the threshold (>6METs) for vigorous activity. Similarly, short bursts of vigorous exercise, for example the choice to take the stairs, can meaningfully increase average intensity even though they may add only moments to exercise time.

### Novelty and relevance

#### What is new?

- Strong evidence from a large population study that increasing exercise intensity reduces hypertension incidence
- Approach using average habitual exercise intensity including both LTPA and incidental walking and stairclimbing for a more accurate overall assessment of PA

#### What is relevant?

- Intensity of PA has been isolated as study factor by controlling for EE
- Increasing PA intensity is associated with greater protective effect against hypertension than increasing PA time

Clinical/pathophysiological implications:

- Normotensive adults should aim to increase the intensity of their habitual PA to reduce risk of hypertension incidence
- It is more important to increase exercise intensity than to increase or maintain exercise time
- The intensity of habitual PA can be increased without reliance on LTPA: short bursts of intense PA (e.g., stairclimbing) and consistent minor changes (e.g., incrementally increasing walking pace) will also increase average intensity.

## Data Availability

The data underlying this article will be shared on reasonable request to the corresponding author.

## Non-standard abbreviations and acronyms

PA: Physical activity
EE: Energy expenditure
SUN: Seguimiento Universidad de Navarra
EICR: Exercise induced cardiac remodelling
CRF: Cardiorespiratory fitness

## (7) ACKNOWLEDGEMENTS

We especially thank all participants in the SUN cohort for their long-standing and enthusiastic collaboration and our advisors from Harvard TH Chan School of Public Health (Walter Willett, Alberto Ascherio, and Frank B. Hu) who helped us to design the SUN Project. We are also grateful to the other members of the SUN Group for administrative, technical, and material support.

## REFERENCES

1. Fuchs FD, Whelton PK. High blood pressure and cardiovascular disease. Hypertension. 2020;75:285–292.

2. Yusuf S, Joseph P, Rangarajan S, Islam S, Mente A, Hystad P, Brauer M, Kutty VR, Gupta R, Wielgosz A. Modifiable risk factors, cardiovascular disease, and mortality in 155 722 individuals from 21 high-income, middle-income, and low-income countries (PURE): a prospective cohort study. The Lancet. 2020;395:795–808.

3. Murray CJ, Aravkin AY, Zheng P, Abbafati C, Abbas KM, Abbasi-Kangevari M, Abd-Allah F, Abdelalim A, Abdollahi M, Abdollahpour I. Global burden of 87 risk factors in 204 countries and territories, 1990–2019: a systematic analysis for the Global Burden of Disease Study 2019. The Lancet. 2020;396:1223–1249.

4. Gaziano TA, Bitton A, Anand S, Weinstein MC. The global cost of nonoptimal blood pressure. Journal of Hypertension. 2009;27:1472–1477.

5. Mills KT, Stefanescu A, He J. The global epidemiology of hypertension. Nature Reviews Nephrology. 2020;16:223–237.

6. Yusuf S, Hawken S, Ôunpuu S, Dans T, Avezum A, Lanas F, McQueen M, Budaj A, Pais P, Varigos J. Effect of potentially modifiable risk factors associated with myocardial infarction in 52 countries (the INTERHEART study): case-control study. The Lancet. 2004;364:937–952.

7. Lee DH, Rezende LF, Joh H-K, Keum N, Ferrari G, Rey-Lopez JP, Rimm EB, Tabung FK, Giovannucci EL. Long-term leisure-time physical activity intensity and all-cause and cause-specific mortality: a prospective cohort of US adults. Circulation. 2022;146:523–534.

8. Hayes P, Ferrara A, Keating A, McKnight K, O’Regan A. Physical activity and hypertension. Reviews in Cardiovascular Medicine. 2022;23:302.

9. Diaz KM, Shimbo D. Physical activity and the prevention of hypertension. Current Hypertension Reports. 2013;15:659–668.

10. Warburton DE, Nicol CW, Bredin SS. Health benefits of physical activity: the evidence. Canadian Medical Association Journal. 2006;174:801–809.

11. Liu X, Zhang D, Liu Y, Sun X, Han C, Wang B, Ren Y, Zhou J, Zhao Y, Shi Y. Dose–response association between physical activity and incident hypertension: a systematic review and meta-analysis of cohort studies. Hypertension. 2017;69:813–820.

12. Pescatello LS, Buchner DM, Jakicic JM, Powell KE, Kraus WE, Bloodgood B, Campbell WW, Dietz S, DiPietro L, George SM. Physical activity to prevent and treat hypertension: a systematic review. Medicine & Science in Sports & Exercise. 2019;51:1314–1323.

13. Garber CE, Blissmer B, Deschenes MR, Franklin BA, Lamonte MJ, Lee I-M, Nieman DC, Swain DP. Quantity and quality of exercise for developing and maintaining cardiorespiratory, musculoskeletal, and neuromotor fitness in apparently healthy adults: guidance for prescribing exercise. Medicine and Science in Sports and Exercise. 2011;43:1334–1359.

14. World Health Organization. WHO guidelines on physical activity and sedentary behaviour. Geneva: World Health Organisation; 2020.

15. American College of Sports Medicine. ACSM’s guidelines for exercise testing and prescription. Eleventh edition. Philadelphia: Wolters Kluwer; 2021.

16. O’Donovan G, Blazevich AJ, Boreham C, Cooper AR, Crank H, Ekelund U, Fox KR, Gately P, Giles-Corti B, Gill JM. The ABC of Physical Activity for Health: A consensus statement from the British Association of Sport and Exercise Sciences. Journal of Sports Science. 2010;28:573–591.

17. Hidalgo-Santamaria M, Bes-Rastrollo M, Martinez-Gonzalez MA, Moreno-Galarraga L, Ruiz-Canela M, Fernandez-Montero A. Physical activity intensity and cardiovascular disease prevention—from the Seguimiento Universidad De Navarra study. The American Journal of Cardiology. 2018;122:1871–1878.

18. Hidalgo-Santamaria M, Fernandez-Montero A, Martinez-Gonzalez MA, Moreno-Galarraga L, Sanchez-Villegas A, Barrio-Lopez MT, Bes-Rastrollo M. Exercise intensity and incidence of metabolic syndrome: the SUN Project. American Journal of Preventive Medicine. 2017;52:95–101.

19. Llavero-Valero M, Escalada-San Martín J, Martínez-González MA, Basterra-Gortari FJ, Gea A, Bes-Rastrollo M. Physical activity intensity and type 2 diabetes: Isotemporal substitution models in the “Seguimiento universidad de navarra”(SUN) cohort. Journal of Clinical Medicine. 2021;10:2744.

20. Etzig C, Gea A, Martinez-Gonzalez MA, Sullivan Jr MF, Sullivan E, Bes-Rastrollo M. The association between self-perceived walking pace with the incidence of hypertension: the ‘Seguimiento Universidad de Navarra’cohort. Journal of Hypertension. 2021;39:1188–1194.

21. Carlos S, De La Fuente-Arrillaga C, Bes-Rastrollo M, Razquin C, Rico-Campà A, Martínez-González MA, Ruiz-Canela M. Mediterranean diet and health outcomes in the SUN cohort. Nutrients. 2018;10:439.

22. World Medical Association. World Medical Association Declaration of Helsinki: ethical principles for medical research involving human subjects. Journal of the American Medical Association. 2013;310:2191–2194.

23. Willett W. Nutritional epidemiology. 3rd ed. New York: Oxford University Press; 2013.

24. Williams B, Mancia G, Spiering W, Agabiti Rosei E, Azizi M, Burnier M, Clement DL, Coca A, De Simone G, Dominiczak A. 2018 ESC/ESH Guidelines for the management of arterial hypertension: The Task Force for the management of arterial hypertension of the European Society of Cardiology (ESC) and the European Society of Hypertension (ESH). European Heart Journal. 2018;39:3021–3104.

25. Martínez-González MA, López-Fontana C, Varo JJ, Sánchez-Villegas A, Martinez JA. Validation of the Spanish version of the physical activity questionnaire used in the Nurses’ Health Study and the Health Professionals’ Follow-up Study. Public Health Nutrition. 2005;8:920–927.

26. Ainsworth BE, Haskell WL, Herrmann SD, Meckes N, Bassett Jr DR, Tudor-Locke C, Greer JL, Vezina J, Whitt-Glover MC, Leon AS. 2011 Compendium of Physical Activities: a second update of codes and MET values. Medicine & Science in Sports & Exercise. 2011;43:1575–1581.

27. Ainsworth BE, Haskell WL, Herrmann SD, Meckes N, Bassett Jr DR, Tudor-Locke C, Greer JL, Vezina J, Whitt-Glover MC, Leon AS. The Compendium of Physical Activities Tracking Guide. Healthy Lifestyles Research Center, College of Nursing & Health Innovation, Arizona State University https://sites.google.com/site/compendiumofphysicalactivities/. 2011. Accessed September.

28. Alonso A, Beunza JJ, Delgado-Rodríguez M, Martínez-González MA. Validation of self reported diagnosis of hypertension in a cohort of university graduates in Spain. BMC Public Health. 2005;5:1–7.

29. Trichopoulou A, Costacou T, Bamia C, Trichopoulos D. Adherence to a Mediterranean diet and survival in a Greek population. New England Journal of Medicine. 2003;348:2599–2608.

30. Hirshkowitz M, Whiton K, Albert SM, Alessi C, Bruni O, DonCarlos L, Hazen N, Herman J, Katz ES, Kheirandish-Gozal L. National Sleep Foundation’s sleep time duration recommendations: methodology and results summary. Sleep Health. 2015;1:40–43.

31. Swain DP, Franklin BA. Comparison of cardioprotective benefits of vigorous versus moderate intensity aerobic exercise. The American Journal of Cardiology. 2006;97:141–147.

32. Huai P, Xun H, Reilly KH, Wang Y, Ma W, Xi B. Physical activity and risk of hypertension: a meta-analysis of prospective cohort studies. Hypertension. 2013;62:1021–1026.

33. Lee LL, Mulvaney CA, Wong YKY, Chan ES, Watson MC, Lin HH. Walking for hypertension. Cochrane Database of Systematic Reviews. John Wiley & Sons, Ltd; 2021.

34. Igarashi Y, Nogami Y. Running to lower resting blood pressure: a systematic review and meta-analysis. Sports Medicine. 2020;50:531–541.

35. Paffenbarger Jr RS, Wing AL, Hyde RT, Jung DL. Physical activity and incidence of hypertension in college alumni. American Journal of Epidemiology. 1983;117:245–257.

36. Williams PT, Thompson PD. Walking versus running for hypertension, cholesterol, and diabetes mellitus risk reduction. Arteriosclerosis, thrombosis, and vascular biology. 2013;33:1085–1091.

37. Pavey TG, Peeters G, Bauman AE, Brown WJ. Does vigorous physical activity provide additional benefits beyond those of moderate? Medicine and Science in Sports and Exercise. 2013;45:1948–1955.

38. Wilson M, Ellison GM, Cable NT. Basic science behind the cardiovascular benefits of exercise. Heart. 2015;101:758–765.

39. Swain DP, Franklin BA. VO(2) reserve and the minimal intensity for improving cardiorespiratory fitness. Medicine and Science in Sports and Exercise. 2002;34:152–157.

40. Cheng C, Zhang D, Chen S, Duan G. The association of cardiorespiratory fitness and the risk of hypertension: a systematic review and dose–response meta-analysis. Journal of Human Hypertension. 2022;36:744–752.

41. Weiner RB, Baggish AL. Exercise-induced cardiac remodeling. Progress in Cardiovascular Diseases. 2012;54:380–386.

42. Levine BD. Can intensive exercise harm the heart? The benefits of competitive endurance training for cardiovascular structure and function. Circulation. 2014;130:987–991.

43. Nes BM, Gutvik CR, Lavie CJ, Nauman J, Wisløff U. Personalized activity intelligence (PAI) for prevention of cardiovascular disease and promotion of physical activity. The American Journal of Medicine. 2017;130:328–336.

